# Kolmogorov-Arnold Network for Atherosclerotic Cardiovascular Disease Risk Prediction

**DOI:** 10.1101/2025.05.25.25328329

**Authors:** Sang-Kyun Ko, Sun-Wha Kim, Hwi Kwon, Si-Hyuck Kang, Chang-Hwan Yoon, Tae-Jin Youn, In-Ho Chae

**Author notes:** Corresponding author *Email address:* (Si-Hyuck Kang).

## Abstract

**Background:** Assessing the risk of future atherosclerotic cardiovascular disease (ASCVD) is crucial in clinical practice, yet it continues to pose significant challenges in cardiology. The Kolmogorov–Arnold Network (KAN) has recently been proposed as an alternative to traditional multi-layer perceptrons (MLP). This study aims to develop a 10-year ASCVD risk prediction model using KAN and to compare its performance with pre-existing and other machine learning-based techniques.

**Methods:** We utilized data from 2,116,621 individuals in the Korean National Health Insurance Service cohort for training and validating the models. Alongside KAN, we developed models using MLP, logistic regression, and random forest. Conventional equation models included PREVENT, PCE, Korean Risk Prediction Model, and SCORE2. We evaluated the models using the area under the receiver operating characteristic curve (AUROC) and the area under the precision–recall curve (AUPRC).

**Findings:** The KAN-based model achieved an AUROC of 0.7765 (95% confidence intervals, 0.7461-0.8068) and an AUPRC of 0.1551, outperforming both conventional equation-based models and other machine learning approaches. KAN offered interpretability by enabling the extraction of symbolic formulas and visualizing the contributions of individual risk factors through pruned network graphs. We further assessed the influence of risk factors on ASCVD prediction using SHAP analysis, ablation studies, and model output visualizations.

**Interpretations:** The KAN-based model demonstrated superior performance and enhanced interpretability in predicting ASCVD risk. These findings suggest that KAN could be a promising alternative to other machine learning models in medical applications.

**Funding:** This research was supported by the Bio&Medical Technology Development Program of the National Research Foundation (NRF) funded by the Korean government (MSIT) (NO. RS-2023-00222910) (RS-2025-00517929).

**Copyright:** This preprint is made available under a CC BY-NC 4.0 license. Copyright remains with the author(s).

**Research in context:** *Evidence before this study:* Predicting 10-year atherosclerotic cardiovascular disease (ASCVD) risk is a clinically important task in cardiology. In recent years, numerous studies have shown that machine learning (ML) models outperform conventional equation-based risk prediction models such as the Pooled Cohort Equations (PCE), PREVENT, and SCORE2. However, widely used ML models such as multilayer perceptrons (MLPs) often lack interpretability and involve complex architectures, which makes them difficult to implement in clinical settings. To investigate whether the recently proposed Kolmogorov–Arnold Network (KAN) could offer a more interpretable machine learning–based alternative in 10-year ASCVD task, we conducted a comprehensive literature search up to April 2025 using PubMed, Google Scholar, Open-Review, and arXiv. Search terms included “atherosclerotic cardiovascular disease”, “risk prediction”, “Kolmogorov-Arnold Network”, “machine learning”, “deep learning”, “multilayer perceptrons”, “cardiovascular disease”, and “medical data”. Despite the growing interest in ML–based cardiovascular risk prediction, we found no studies, that applied KAN to ASCVD risk estimation. Existing studies using KAN in cardiology have focused on tasks such as synthetic ECG signal analysis or classification, but not long-term ASCVD risk prediction. This highlights a clear gap in the literature for interpretable machine learning models that can provide equation-like outputs in real-world population-based risk prediction.

*Added value of this study:* Although multilayer perceptrons (MLPs) have shown strong performance in predicting 10-year ASCVD risk, their lack of interpretability limits clinical adoption, where conventional models like PCE and PREVENT remain dominant. In this study, we propose a Kolmogorov–Arnold Network (KAN)–based model that generates simple and interpretable equations while maintaining high predictive accuracy. Using over 2.1 million individuals from the Korean National Health Insurance Service cohort with 10-year follow-up, the KAN model outperformed both traditional equation models and other ML approaches in AUROC and AUPRC. To our knowledge, this is the first study to apply KAN to long-term ASCVD risk prediction, bridging the gap between interpretability and machine learning performance in a clinically meaningful context.

*Implications of all the available evidence:* The growing body of evidence supports the use of ML models for predicting future ASCVD risk, yet lack of interpretability makes most models remain impractical for clinical use. Our findings demonstrate that interpretable ML approaches such as KAN can bridge this gap. By producing symbolic formulas with competitive performance, the model may enable clinicians to adopt ML-driven risk scores within familiar clinical workflows. Broader adoption of such approaches could support personalized prevention strategies and improved risk communication in cardiovascular care. We also hope that our study serves not only as a proposal of a novel predictive model, but also as a foundation that inspires future research on clinically applicable prediction models using KAN, further expanding its utility beyond theoretical development.

## Introduction

Cardiovascular disease (CVD) is the leading cause of death, accounting for one third of global deaths.^1^ Previous studies have suggested that the development of atherosclerotic CVD (ASCVD) can be predicted based on risk factors with a reasonable degree of precision. Contemporary clinical guidelines have adopted individual assessment of future ASCVD risk as the key step in clinical decisions such as initiation of lipid-lowering,^2,3^ or blood pressure (BP) lowering medical therapy.^4,5^ The models include the Framingham Heart Study,^6^ the SCORE project for the European population,^7,8^ the Pooled Cohort Equations (PCE) for White and African American populations in the United States,^9^ the QRISK study for the UK population,^10,11,12^ and the PREVENT study.^13^ All the mentioned models provide a function that estimates the risk of ASCVD over the next 10 years, and use a small number of variables, typically between 6 and 13, making them practical for clinical use. Newer models have been suggested over recent decades to provide greater precision and accuracy and support a wider population.

Over the past decade, advances in hardware capable of parallel computation have made machine learning theories possible, and the field has demonstrated outstanding performance in solving many difficult problems. The model known as the feedforward neural network, or multi-layer perceptron (MLP), has been widely used in most architectures and is considered one of the most successful models.^14^ We previously reported that an MLP-based model provides better discrimination and higher calibration than preexisting cardiovascular risk prediction models. Other studies also demonstrated that MLP-based risk estimation achieved high AUROC values.^15^ However, the MLP commonly used for ASCVD tabular data makes it challenging to understand the meaning of each parameter and how they contribute to specific outcomes.^16^ The low interpretability and high complexity of MLP, even if it achieves better performance than models such as those referenced in,^17,18,7,8,13,6,9,10,11,12^ can reduce its potential for clinical application in ASCVD risk prediction.

Recently, the Kolmogorov-Arnold Network (KAN)^19^ emerged as a potential alternative to MLP. The Universal Approximation Theorem^20^ ensures that MLPs can approximate arbitrary non-linear functions under certain conditions. Unlike MLPs, KAN is based on the Kolmogorov–Arnold representation theorem,^21,22,23^ which may offer greater expressiveness and improved accuracy with a smaller model size. KAN replaces densely connected nodes with trainable activation functions assigned to weights, facilitating easier understanding and interpretation of complex expressions. Furthermore, other studies have empirically demonstrated that KAN can outperform MLP in both accuracy and computational efficiency for specific tasks.^24,25^ In contrast, Somvanshi et al.^26^ report that KAN requires further improvements in computational efficiency when dealing with high-dimensional and noisy data. While additional research is needed to determine whether KAN consistently outperforms MLP, our focus lies not in maximizing performance, but in leveraging the interpretability potential of KAN. Interpretability is another strength of KAN that can address the ‘black box’ problem in medical AI. As their architecture uses learnable univariate activation functions that enable symbolic representation and pruning, KANs can be intuitively visualized and can easily interact with human users. In this context, our study focuses on the strong expressiveness of the KAN model and aims to extract meaningful mathematical expressions from a trained network for ASCVD risk prediction.

Therefore, in this study, we built a model predicting 10-year ASCVD risk using KAN architecture. We compared its discrimination and performance with those of pre-existing equation models and other machine learning approaches. We also showed how the proposed function trained through KAN provides an interpretable function as shown in previous studies.^7,8,13,6,9,10,11,12^

## Related works

Established risk estimation models typically adopted linear regression models with a varying number of baseline risk factors and a small variation in the definition of ASCVD. Equation models evaluated and validated in this study include PCE, SCORE2, KRPM, and PREVENT. In 2013, the American College of Cardiology (ACC) and the American Heart Association (AHA) jointly introduced the Pooled Cohort Equations (PCE), a 10-year ASCVD risk prediction model based on data from White and Black populations in the United States.^9^ Variables include age, systolic blood pressure (treated or untreated), total cholesterol, HDL cholesterol, diabetes status, and current smoking status as predictive variables. The SCORE model was introduced in 2003 by the European Society of Cardiology and was designed to predict only CVD mortality. SCORE2 is a newer algorithm that estimates 10-year risk of fatal and nonfatal cardiovascular disease (CVD) events in individuals aged 40-69.^8^ The model utilizes data from 677,684 individuals across 45 cohorts in 13 European countries, with 30,121 CVD events analyzed. The SCORE2 equation model employs the fewest variables among the models considered in this study, including sex, age, systolic blood pressure, total cholesterol, HDL cholesterol, smoking status, and diabetes status. A Korean-specific model, Korean Risk Prediction Model (KRPM), was developed in 2015 utilizing data from 200,010 Koreans aged 40–79 based on the Korea Heart Study cohort.^27^ The included variables and the model development scheme were similar to those of the PCE. PREVENT is the most up-to-date risk prediction model endorsed by the American Heart Association.^13^ The novel equation is sex-specific and race-free, and predicts risk of total cardiovascular disease including ASCVD and heart failure. The base model additionally includes estimated glomerular filtration rate, and add-on models offer the flexibility to include additional measures of kidney (urine albumin-to-creatinine ratio), metabolic (hemoglobin A1c), and social (social deprivation index) risk.

Studies have suggested that machine learning approaches for estimating ASCVD risk showed higher performance than traditional equation-based risk models. Alaa et al. (2019)^28^ demonstrated that machine learning methods, applied to UK Biobank data from ~ 400, 000 individuals, outperformed traditional equation-based risk models like Framingham and QRISK while also identifying novel predictive variables for cardiovascular disease risk. Li et al.^29^ utilized electronic health record (EHR) data from 1.1 million individuals in the UK (1985–2015) to predict ASCVD risk, applying traditional risk models, machine learning methods, and the transformer-based BEHRT architecture. Unlike conventional approaches, their study treated data from the Clinical Practice Research Datalink (CPRD) as a time series, enabling BEHRT to process sequential medical information, including diagnoses, medications, lab tests, and procedures. BEHRT outperformed equation-based models such as QRISK3, Framingham, and ASSIGN, as well as the Random Forest algorithm, achieving the highest predictive accuracy for heart failure, stroke, and coronary artery disease risk. Mamouei et al.^30^ demonstrated that BEHRT outperformed traditional models, including Random Forest, in analyzing approximately 500,000 records from the UK Biobank. Although its performance declined during external validation with a time shift, BEHRT still achieved the highest accuracy under temporal data shift conditions.^29^ There are also many other studies that demonstrated improved predictive performance of machine learning-based methods.^15,18,31,32^

Korean ethnicity was poorly represented in the development stage of representative ASCVD risk prediction models. Cho et al.^33^ analyzed approximately 500,000 Korean subjects from the National Health Insurance Service-National Health Screening Cohort to show that Framingham risk score, SCORE, and QRISK2 overestimated risk while PCE demonstrated good calibration. A Korean-specific risk prediction model (KRPM) for ASCVD^27^ has been developed using data of 200,100 subjects from the Korean Heart Study cohort.^34^ Researchers have developed various ASCVD prediction models for Koreans using different machine learning approaches.^35,36,37^

KAN,^19^ which was released in 2024, provoked a rapid surge of related research to verify its improved performance and interpretability.^38,39,40,41^ In this section, we briefly review recent studies in the field of cardiology. Jahin, M. A. et al.^42^ proposed Kolmogorov-Arnold Classical-Quantum Dual-Channel Neural Network trained on the public Heart failure prediction dataset,^43^ which achieved predictive performance that surpassed 34 other benchmark models. Zhang et al. integrates optimal transport, structure remapping, and KAN into a graph neural networks-based model to evaluate combined cardiotoxicity on cardiac ion channels.^44^ Huang et al. used KANs to analyze electrocardiogram signals in order to detect cardiac abnormalities.^45^ Zeleke et al. incorporated KAN into a federated learning framework and demonstrated significant performance improvements on both real-world and synthetic electrocardiogram datasets.^46^ However, attempts have yet to be made to derive a function to predict ASCVD risk using KAN.

## Methods

### Data source and study population

This is a retrospective study of a nationwide healthcare utilization database, the National Health Insurance Service (NHIS) Cohort, Republic of Korea. Details about the cohort have been described previously.^47^ A total of 2,854,149 individuals who underwent a general health screening program between 2009 and 2012 were extracted. Selection criteria included (1) age between 30 and 79 years, and (2) no previous diagnosis of CVDs, such as myocardial infarction, ischemic stroke, and congestive heart failure during the previous 2 years. (3) Those with angina who received coronary revascularization therapy, such as percutaneous coronary intervention and coronary artery bypass surgery were excluded. (4) In addition, to avoid bias caused by statin therapy, individuals who had been receiving a statin before the screening or started statin therapy during the study period before the obtaining of the study outcomes were also excluded. The final study population consisted of 2,116,621 individuals.

This study was exempt from review by the Seoul National University, Bundang Hospital Institutional Review Board (I-2018-11316). It complied with the requirements of the Declaration of Helsinki, and the need for informed consent was waived.

### Definition of independent variables

Twenty two variables were selected as risk factors as shown in Table 1. Eight variables that were used in the established risk prediction models were considered in the primary analysis model, while an additional 14 variables were included in the extended model. Notably, PREVENT additionally includes estimated glomerular filtration rate (eGFR) and body mass index (BMI) as a continuous variable, as well as urine albumin creatinine ratio, HbA1c, and social deprivation index (SDI) as optional variables. Unfortunately, HbA1c was not available in this cohort, a categorical variable of urine protein colorimetric dipstick test (ie. absent, trace, 1+, 2+, 3+, and 4+) was used instead of urine albumin creatinine ratio, and Zip code was instead of social deprivation index. Other variables included in the extended model were diastolic BP, triglycerides, hemoglobin, aspartate aminotransferase, alanine aminotransferase, gamma-glutamyl transferase, fasting blood sugar, low-density lipoprotein, waist circumference, which were routinely measured during the health screening program. We assume these tests are within routine clinical practice recommended by contemporary guidelines.^4,48^

**Table 1:**
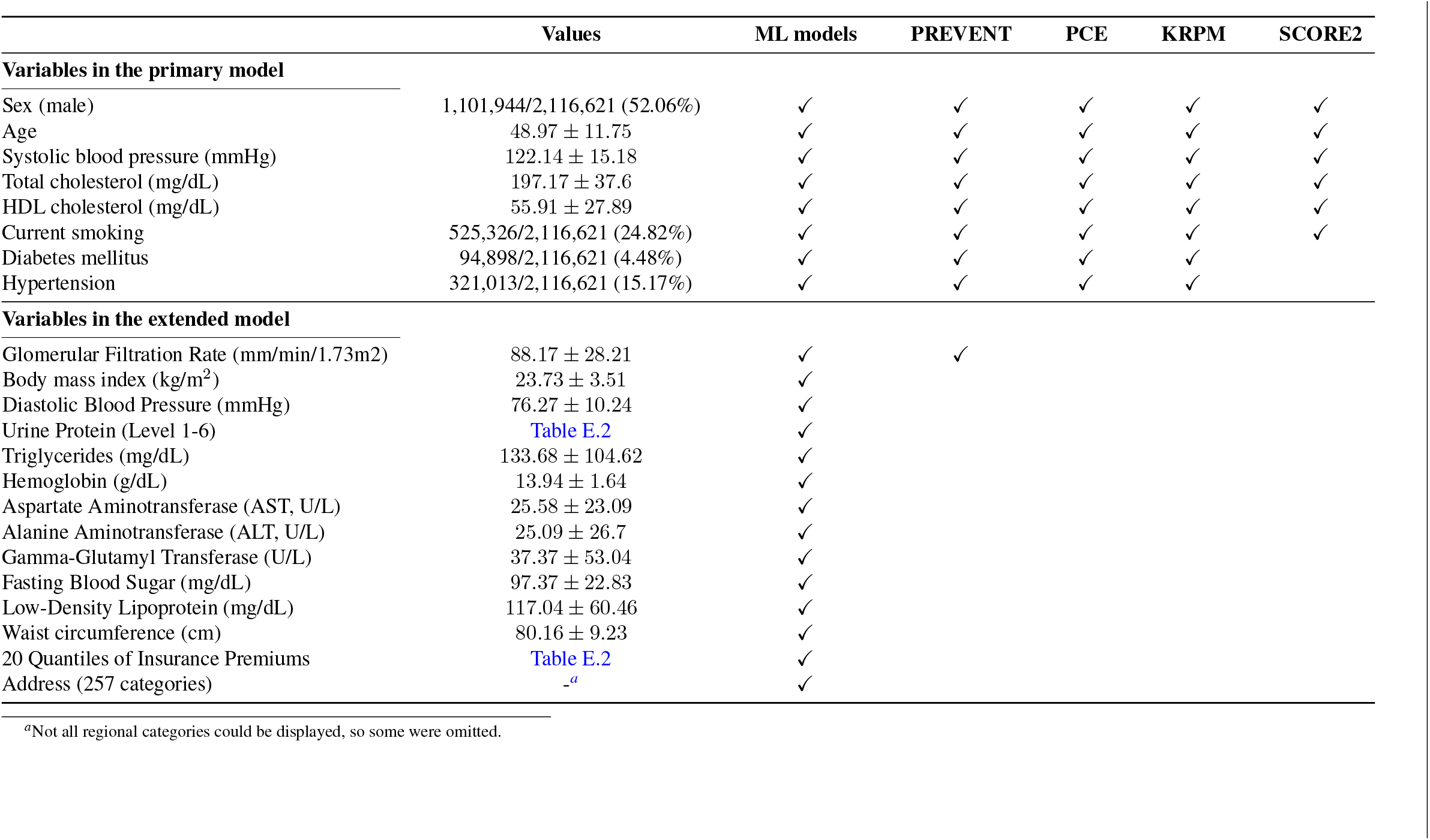
Baseline characteristics of the study population and predictor variables used in each model. Eight risk factors listed in the upper panel are included in the primary model, while 14 variables in the lower panel are added in the extended model.

Demographic characteristics were extracted from the enrollment status database. Histories of diabetes and hypertension medication use were identified using previous claims data during the previous year. Smoking status and the amount of smoking were identified using self-report questionnaires. Results of anthropometric and blood pressure (BP) measurements, and laboratory tests using blood and urine samples were obtained from the health screening program. Missing values of eGFR were imputed using the single imputation method. Regression imputation strategy was used: as eGFR exhibited a negative linear relationship with age, missing values were imputed by calculating the mean eGFR within specific age groups, stratified by gender. We then applied outlier filtering to the data based on the criteria described in Table E.1.

### Definition of dependent variables

The study clinical endpoint was defined as ASCVD, a composite of cardiac death, myocardial infarction, and ischemic stroke. Recently, the PREVENT equation adopted a total CVD that includes HF event as well as fatal and nonfatal ASCVD.^13^ However, HF was not included in this study because risk of ASCVD is yet considered the primary factor for treatment decision according to most clinical practice guidelines.^2,49^ Definition of each event was described previously.^33^

## Models

### Machine learning models

The data **x** ∈ ℝ *D*, consisting of *D*-dimension health examination variables, corresponds to a target value *y* ∈ {0, 1}, where 0 or 1 indicates whether ASCVD occurs within 10-years. The predictor vector **x** includes various combinations of features as defined in Table 1. The dataset *D* consists of *N* individual health examination data and corresponding ASCVD outcomes, represented as pairs (**x**, *y*), and is defined as 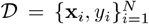. The prediction model for ASCVD risk can be defined as *f* : ℝ^*D*^ → [0, 1]^2^, returning two real numbers that sum to 1. Therefore the predicted value *f* (**x**_*i*_) = *ŷ*_*i*_ is defined as follows:

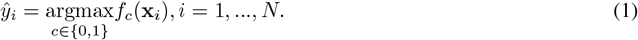

The dataset was split into 80% for training and 20% for testing. Event rates of ASCVD were evenly distributed across the training (72,528 out of 1,588,924 cases, 4.57%) and test sets (18,029 out of 397,232 cases, 4.54%). The training dataset was further split into a training set and a validation set at a 8:2 ratio to tune the hyperparameters of KAN and MLP. The hyperparameter tuning process involved five-fold cross-validation, where model performance was evaluated on each corresponding validation set.^50^ The hyperparameter set that achieved the highest average performance across the five validation sets was chosen for performance evaluation on the test dataset. All training data were scaled using scikit-learn’s `StandardScaler`,^51^ and the test data were transformed using the mean and scale parameters derived from the training set. Further details regarding the scaler are provided in the Appendix B.

### Multi-layer perceptron

The multi-layer perceptron *f* ^MLP^ consists of *L* fully connected layers, each with *h* nodes in the hidden layers. To prevent overfitting to the training data, we incorporated dropout layers^52^ during the construction of the network. We train the model using the Adam optimizer^53^ and mini-batch learning, and implement all components in PyTorch.^54^ Hyperparameter tuning targets include *L, h*, the learning rate, and the dropout rate. We select the hyperparameter set that yields the lowest average loss across five validation sets. To find the optimal combination, we perform an exhaustive search over all possible configurations: we consider *L* ∈ {2, 3, 4}, *h* ∈ {8, 16, 22, 32}, learning rate *∈* {10^−2^, 10^−3^, 10^−4^, 10^−^}^5^, and dropout rate *∈* {0.1, 0.2}. We apply early stopping if the validation loss fails to improve for 50 consecutive steps, with a maximum training limit of 5000 epochs. Detailed mathematical modeling is described in Appendix A.1.

### Kolmogorov-Arnold Network

We train and validate the *f* ^KAN^ using the official code released in December 2024.^55^ Following the example provided in the official implementation, KAN returns two output values corresponding to the binary classification task, as as previously defined. Appendix A.2 provides additional information on KAN, including a detailed example of the two-output version that is not described in the origin paper.^19^ All evaluations of KAN are based not on the network output itself, but on two symbolic formulas obtained after training. The formula corresponding to class 1 is selected for further evaluation. We tune the hyperparameters of KAN according to the guidelines in the ‘Advice on hyperparameter tuning’ section of the official code. We search for an optimal combination of hyperparameters using validation data, with layer depth *∈* {1, 2, 3}, width *∈* {2, 4, 8, 16}, lambda *∈* {10^−2^, 10^−3^, 10^−4^}, and lambda entropy 1, 2, while keeping the grid size and order fixed at 3. The training epoch was determined based on the validation loss history, with a maximum of 5000 epochs. For performance evaluation, we used the second formula extracted by KAN’s symbolic formula function, which predicts the ASCVD class.

### Conventional machine learning models

This study evaluates the performance of the proposed KAN-based risk prediction model in comparison to conventional machine learning methods, using logistic regression and random forest as benchmark models. We selected these two models because logistic regression is a representative linear method, and random forest is a widely used non-linear method. Both are commonly applied as baseline models for performance comparison in the development of 10-year ASCVD risk prediction models. This study implements logistic regression using PyTorch,^54^ following the procedure outlined below: The logistic regression model *f* ^*LR*^ uses a trainable weight vector **w** *∈* ℝ^*D*^ to learn from a *D*-dimensional random variable **x** *∈* ℝ^*D*^. The model returns the output using a non-linear activation function as follows: *f*(**x**) = *σ*(**w**^*T*^ **x**), where *σ* denotes the sigmoid function^1^. The random forest model was trained using the default settings provided by scikit-learn.^56^

### Conventional equation models

PREVENT, PCE, KRPM, and SCORE2 were compared with machine learning models in this study. For fair comparison, the ASCVD risk model of the PREVENT was utilized in this study (not total CVD risk)^13^ (see table Table 1). While the PCE equation is race- and sex-specific, PCE for whites was chosen because it showed the best discrimination and calibration in the Korean population.^9,33^ KRPM is sex-specific and was calculated following the formula.^27^ SCORE2 includes recalibration based on age, sex, and region-specific multipliers; the version for low-risk regions was chosen based on prior findings that it outperformed the high-risk model in Koreans.^8,33^ Conventional equation models were also implemented in Python.

### Experiments

Statistical analysis was done with Python 3.8.16. Model performance was tested in the randomly sampled test dataset. The area under the receiver operating characteristic curve (AUROC) and the area under the precision–recall curve (AUPRC) were calculated using functions from the scikit-learn package.^56^ AUPRC, unlike AUROC, focuses exclusively on the positive class and evaluates the trade-off between precision and recall. AUPRC is considered more informative than AUROC for highly imbalanced classification tasks such as that in this study.^57^

For KAN performance, we extracted the final formula using the `symbolic formula` function provided by the official KAN repository.^19^ We converted the formula into a callable function using the sympy package,^58^ then generated outputs based on this function to evaluate performance. Therefore, the reported metrics represent the performance of an equation derived from the final trained KAN model.

The 95% confidence intervals for the AUROCs of each model shown in Table 2 were calculated using DeLong’s method.^59^ Shapley additive explanations (SHAP) analysis was conducted using the default settings of the `KernelExplainer` function from the Python SHAP package.^60^ An ablation study was performed by removing a single variable from the extended model. To further investigate how the KAN model responds to changes in specific input variables, we generated model output curves that visualize variations in predicted ASCVD risk with respect to the value of a single random variable. Although risk factors do not act independently in real patients, this analysis allows us to observe the model’s sensitivity to each variable in isolation. To generate the model output curves, we used the unique values of a given random variable as the x-axis. Since plotting the output for every individual sample would not be feasible, we calculated the mean KAN output across all test samples sharing the same input value and plotted that mean on the y-axis. For example, if the input variable is age, the x-axis would include all unique values such as 30, 31, 32, and so on. For the value 30, the y-axis would show the average KAN output for all test samples where the age equals 30.

**Table 2:** Area under the receiver operating characteristic (AUROC) and the precision recall curve (AUPRC) for machine learning models compared to established risk prediction models.

| Models | AUROC (95% CI) |  | AUPRC (95% CI) |  |
| --- | --- | --- | --- | --- |
|  | Extended | Primary | Extended | Primary |
| Kolmogorov-Arnold network | **0.7765** (0.7461-0.8068) | **0.7735** (0.7436-0.8033) | **0.1551** | **0.1504** |
| Multi-layer perceptron | 0.7720 (0.7414-0.8026) | 0.7727 (0.7435-0.8020) | 0.1475 | 0.1443 |
| Logistic regression | 0.7714 (0.7413-0.8016) | 0.7720 (0.7426-0.8014) | 0.1505 | 0.1467 |
| Random forest | 0.7489 (0.7195-0.7783) | 0.7049 (0.6771-0.7328) | 0.1403 | 0.0993 |
| PREVENT | 0.7732 (0.7421-0.8043) |  | 0.1541 |  |
| PCE | 0.7680 (0.7357-0.8004) |  | 0.1489 |  |
| KRPM | 0.7730 (0.7440-0.8021) |  | 0.1506 |  |
| SCORE2 | 0.7662 (0.7336-0.7988) |  | 0.1483 |  |

## Results

We compared quantitative performance metrics across four conventional equation models and four machine learning models, including KAN, MLP, logistic regression, and random forest. In this chapter, we go beyond simply reporting predictive performance by extracting symbolic formulas from the KAN model, providing interpretable equations in a form comparable to conventional equation-based models. Furthermore, we explore how and to what extent individual risk factors contribute to ASCVD risk using visual tools such as SHAP analysis, ablation studies, and model output curves.

### Cohort description

Summary statistics of the study population are provided in Table 1. While 52% of the subjects were male, the mean age was 48.9 years. The mean systolic and diastolic BP was 122/76 mmHg, and the levels of total and low-density lipoprotein cholesterol were 197 mg/dL and 117 mg/dL, respectively. The prevalences of hypertension, diabetes, and cigarette smoking were 24.8%, 4.5%, and 15.2%, respectively. Figure D.1 presents a heatmap illustrating the correlations among the variables. Details in categorical variables such as urine protein and income quantiles are provided in Table E.2.

During the study period of 10 years, 96,796 out of 2,116,621 study subjects (4.57%) developed ASCVD. Cardiac death, nonfatal myocardial infarction, and ischemic stroke occurred in 9,298 (0.44%), 31,296 (1.48%), and 54,100 (2.56%) subjects, respectively. Kaplan–Meier survival curves are provided in Figure D.2.

### Model performance

The performance of the study models was compared in the test dataset. Performance of both extended and primary models with a varying number of baseline features was reported for ML models. KAN achieved the highest performance for both sets of baseline features: AUROC for the extended and primary KAN models, 0.7765 (95% CI^2^, 0.773–0.780) and 0.7735 (95% CI, 0.746-0.801), respectively. The primary KAN model exhibited a higher AUROC than all conventional equation models. MLP and logistic regression performed better with the primary model using fewer input variables, whereas KAN and random forest achieved superior results with the extended model that included a larger number of variables. Interestingly, random forest, a machine learning method commonly applied in the medical domain, showed significantly inferior performance.

### Interpretability of KAN

Interpretability is one of the key advantages of KANs. Figure 2 shows the network graph of the KAN model after pruning. Auto-pruning produces a simplified KAN architecture by applying regularization and removing activation functions that contribute very little. Activation functions with values close to zero or minimal contribution appear as disconnected edges. The circles at the bottom represent input variables, and the two circles at the top indicate the ASCVD output. The darkness of each edge reflects its relative importance. Each box along the edge illustrates whether the relationship is linear or nonlinear. As shown in (b) of Figure 2, the variable age is connected by two edges. One of these is the darkest in the graph, and both activation functions show linear patterns. Similarly, smoking status, hypertension, and diastolic blood pressure are connected by relatively dark edges, suggesting stronger influence. In contrast, LDL cholesterol, waist circumference, and eGFR lost their connections after pruning, indicating low relevance to the model output. The network graph of the primary KAN model can be interpreted in the same way.

**Figure 1.**
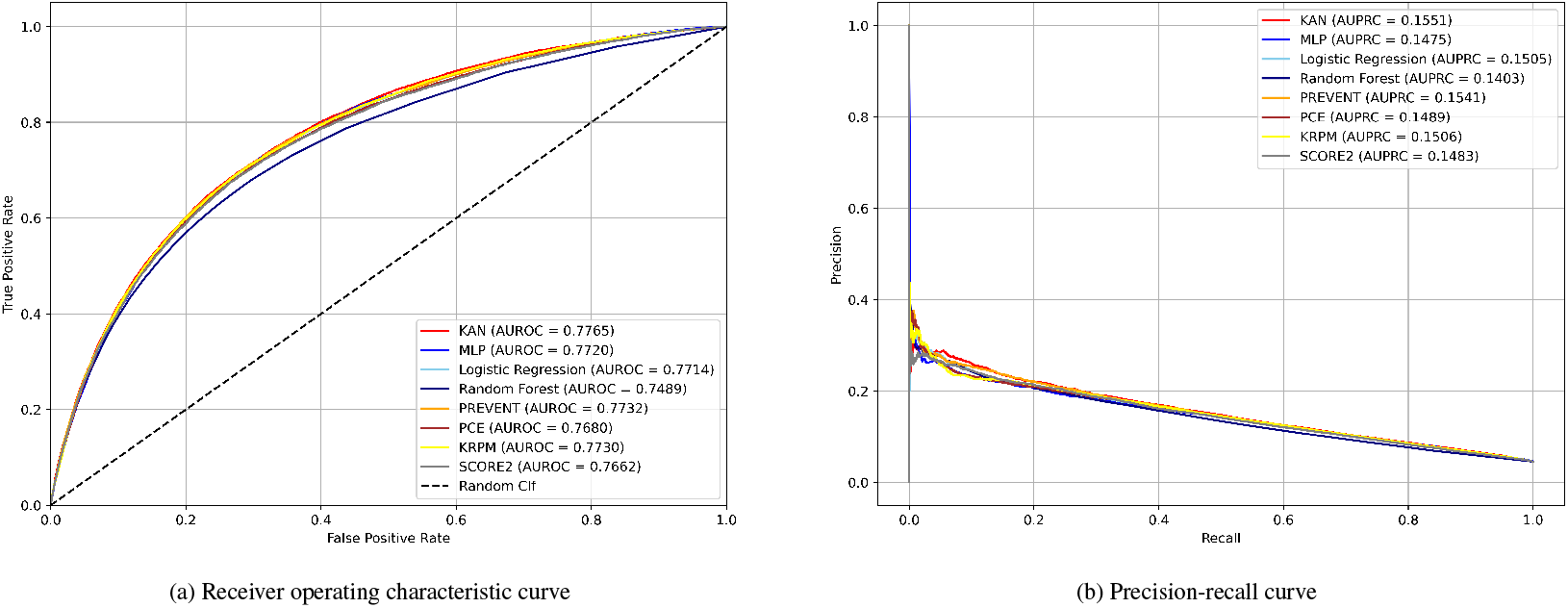
Receiver operating characteristic (ROC) and Precision-Recall curves (PRC) for study models with the extended set of variables. Areas under the ROC (AUROC) and PRC (AUPRC) values are indicated in the legends for comparison.

**Figure 2.**
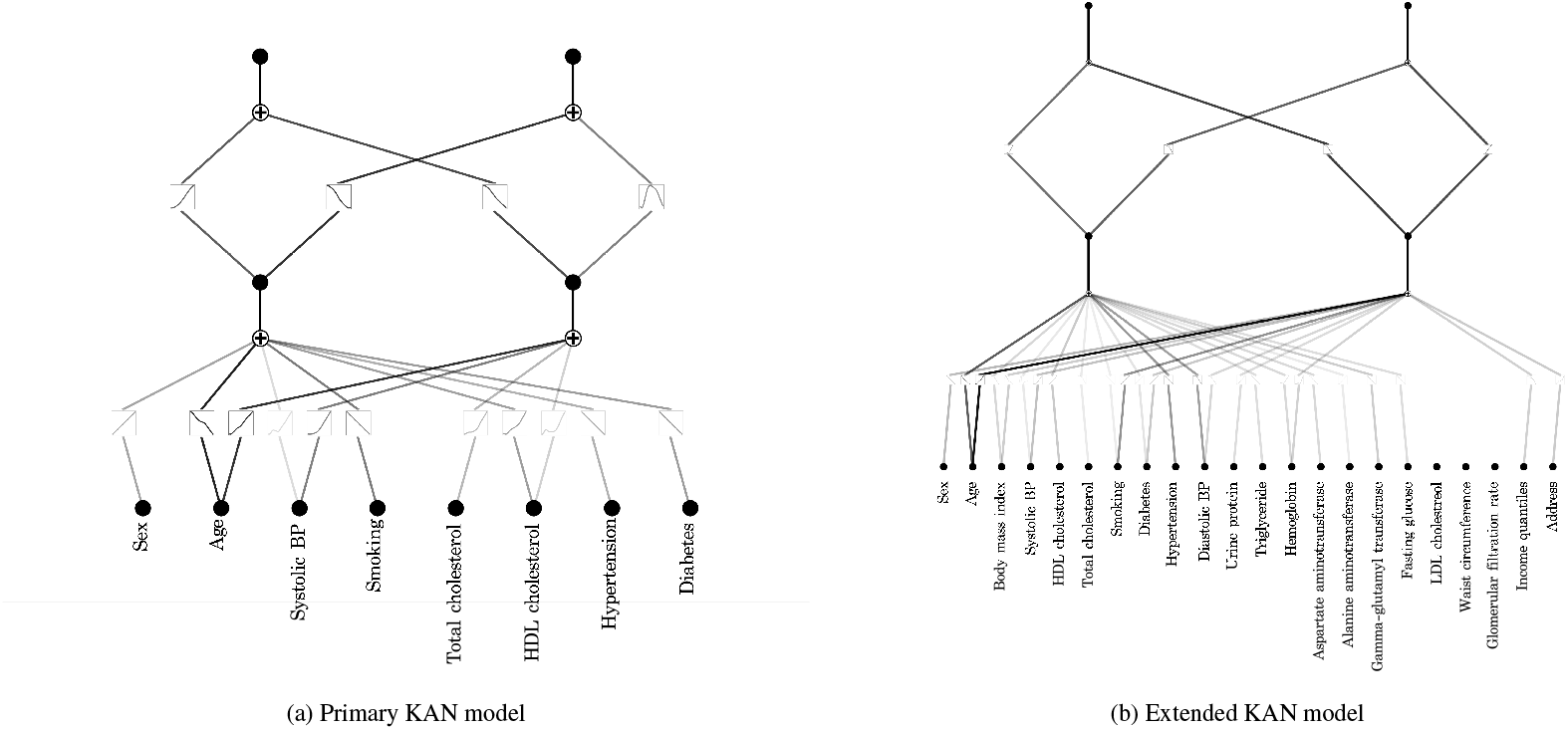
Visualization of KAN after training and pruning.

The KAN model yields a symbolic expression by combining its learned activation functions. The extended KAN model, which demonstrated the best performance in internal test set, is defined as^3^:

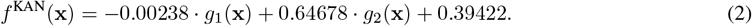

Each function *g*_1_ and *g*_2_ computes the input vector **x** as defined in

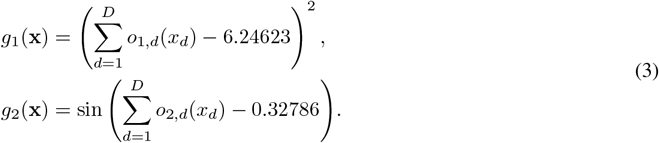

The weights applied to each random variable differ for each *o*_1,*d*_ and *o*_2,*d*_, *d* = 1, …, *D*, and the corresponding weights or operations are described in detail in Table 3.

**Table 3:** List of variables in the extended KAN Equation (2) model with associated weights and operations. All variables were scaled, and the details of the scaling procedure and parameters are provided in the Appendix.

| Risk factors ($x_d$) | $o_{1,d}(\cdot)$ | $o_{2,d}(\cdot)$ |
| --- | --- | --- |
| Sex ($x_1$) | 0 | $-0.08585 \cdot x_1$ |
| Age ($x_2$) | $2.40005 \cdot x_2$ | $0.52258 \cdot x_2$ |
| Body mass index ($x_3$) | $-1.82323 \cdot \sin(0.51184 \cdot x_3 + 8.36848)$ | $0.05827 \cdot x_3$ |
| Systolic BP ($x_4$) | $0.20347 \cdot x_4$ | $0.06788 \cdot x_4$ |
| HDL cholesterol ($x_5$) | $-0.67461 \cdot x_5$ | 0 |
| Total cholesterol ($x_6$) | $0.24987 \cdot x_6$ | 0 |
| Smoking ($x_7$) | $0.24781 \cdot x_7$ | $0.14037 \cdot x_7$ |
| Diabetes ($x_8$) | $0.36863 \cdot x_8$ | $0.06984 \cdot x_8$ |
| Hypertension ($x_9$) | $1.37921 \cdot x_9$ | 0 |
| Diastolic BP ($x_{10}$) | $-5.43350 \cdot \sin(0.40448 \cdot x_{10} - 4.18912)$ | $-0.04471 \cdot x_{10}$ |
| Urine protein ($x_{11}$) | 0 | $0.27216 \cdot \sin(9.98596 \cdot x_{11} + 0.592)$ |
| Triglyceride ($x_{12}$) | $0.44907 \cdot x_{12}$ | 0 |
| Hemoglobin ($x_{13}$) | $-0.47289 \cdot x_{13}$ | $0.03458 \cdot x_{13}$ |
| Aspartate aminotransferase ($x_{14}$) | $0.58114 \cdot x_{14}$ | 0 |
| Alanine aminotransferase ($x_{15}$) | $-0.247468 \cdot x_{15}$ | 0 |
| Gamma-glutamyl transferase ($x_{16}$) | $0.705213 \cdot x_{16}$ | 0 |
| Fasting glucose ($x_{17}$) | $0.3867 \cdot x_{17}$ | 0 |
| LDL cholesterol ($x_{18}$) | 0 | 0 |
| Waist circumference ($x_{19}$) | 0 | 0 |
| Glomerular filtration rate ($x_{20}$) | 0 | 0 |
| Income quantiles ($x_{21}$) | 0 | $-0.055 \cdot x_{21}$ |
| Address ($x_{22}$) | 0 | $0.06013 \cdot x_{22}$ |

### Model interpretation with selected predictors

Figure 3 shows symbolic terms of the extended KAN model output across the range of each input variable. A mostly linear symbolic pattern is shown for age, with predicted risk increasing with age. In contrast, variables such as systolic BP, GFR, LDL-cholesterol, BMI, and waist circumference exhibited nonlinear symbolic terms.

**Figure 3.**
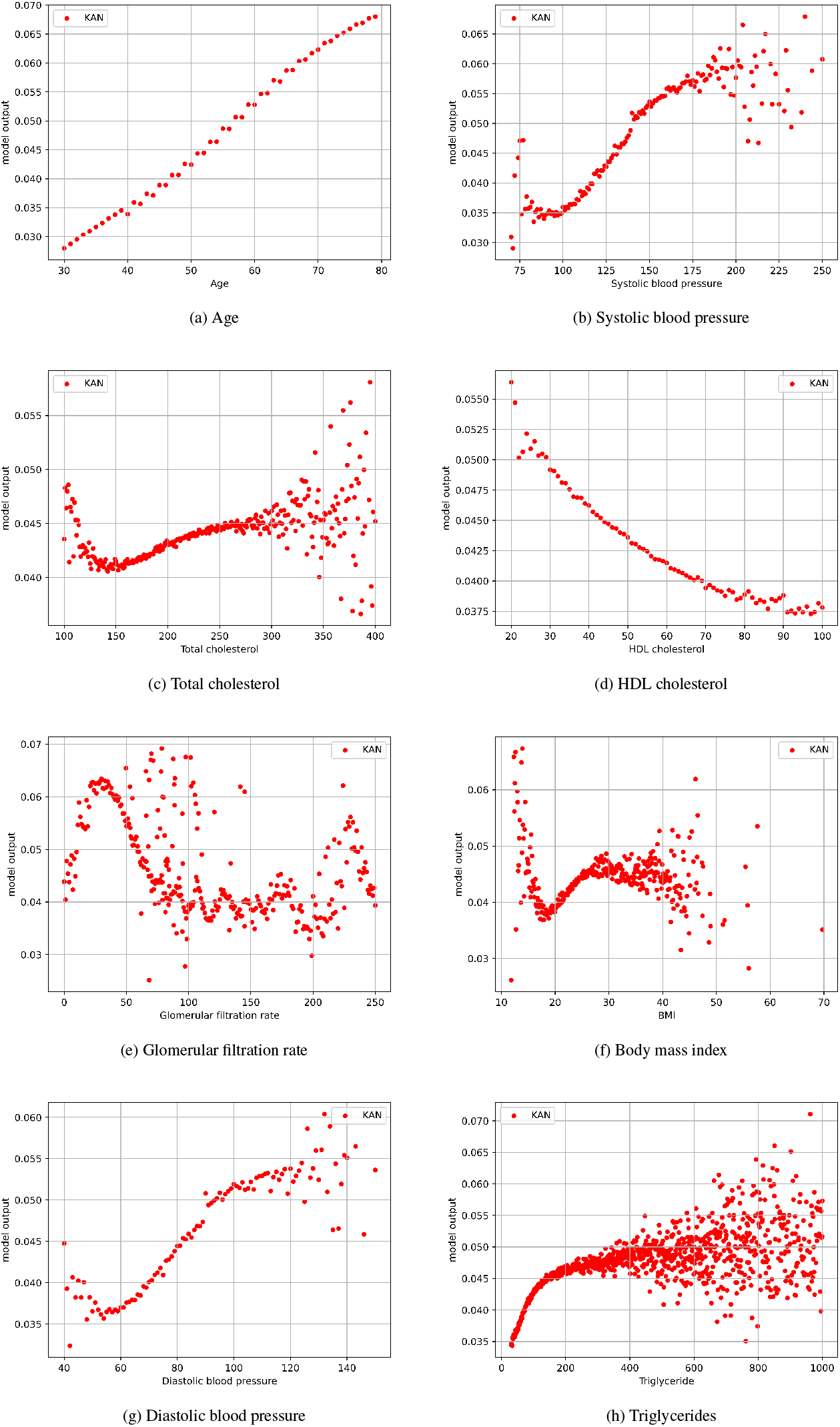
Mean output of the KAN extended model with respect to varying values of each risk factor. Each dot in the scatter plot represents the average prediction across multiple test samples sharing a specific value of the corresponding variable.

The SHAP summary plot illustrates the contribution of each feature to the model’s output Figure 4. A total of 1,000 background data and 1,000 test samples were used. Most of the baseline features showed a linear relationship. Age showed a strong linearity with predicted ASCVD risk. Cigarette smoking and hypertension (red) clearly indicated increased risk. HDL-cholesterol contributed negatively to the output. While BMI exhibited a nonlinear relationship.

**Figure 4.**
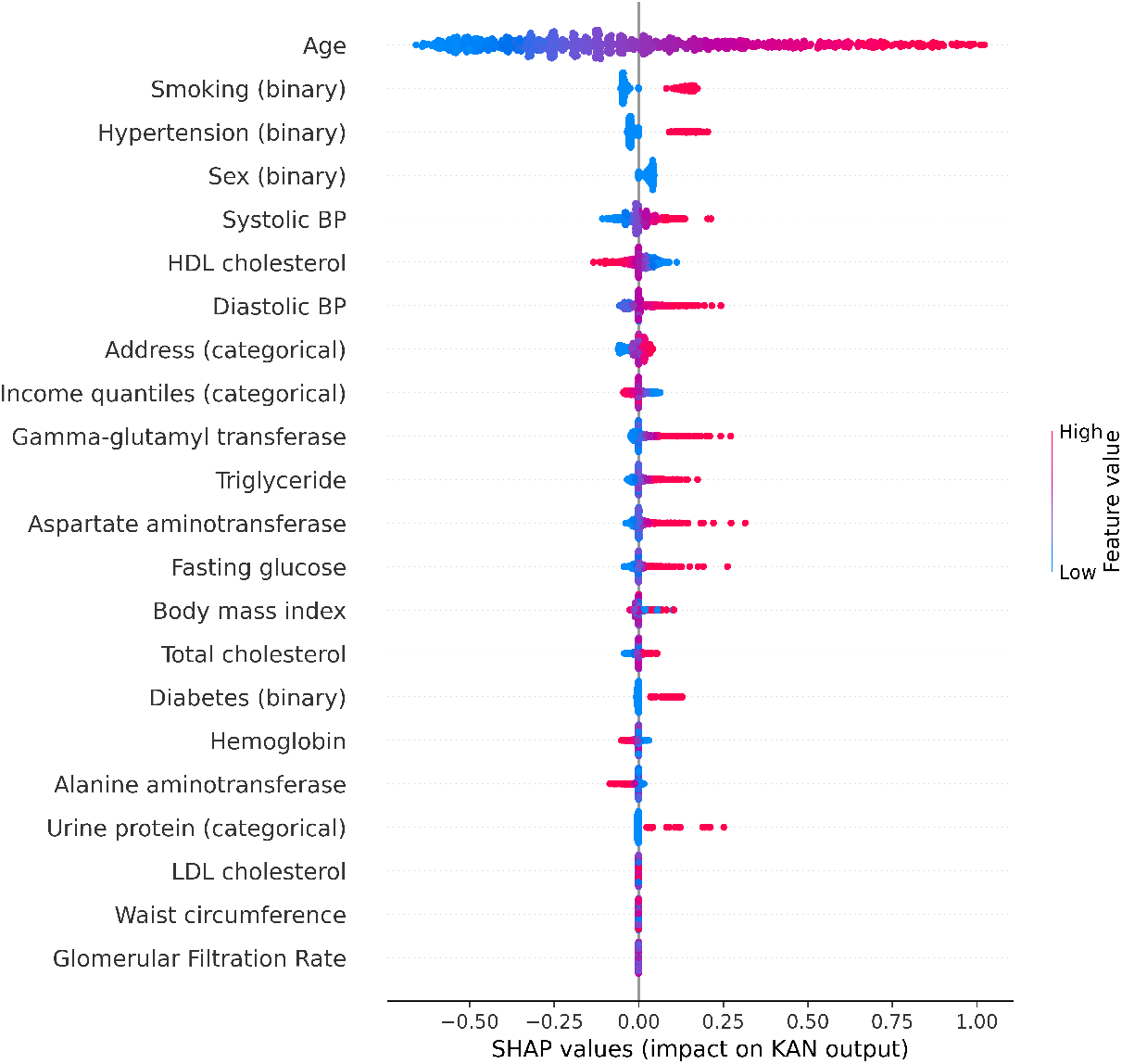
SHapley Additive exPlanations (SHAP) summary plot of 22 variables based on the extended KAN model output. Each point on **the** plot shows a Shapley value for a feature and an instance. The color indicates the value of the feature from blue for low to red for high. The x-axis represents the Shapley value: points with positive SHAP values on the right indicate higher predicted ASCVD risk, while those with negative values on the left do lower risk. The y-axis represents baseline risk factors (feature). A linear association appears when low values (blue points) cluster on the left side and high values (red points) on the right. In contrast, a nonlinear pattern emerges when both low and high values appear on both sides of the axis.

The ablation study in Figure 5 shows contribution of each feature to the performance of the KAN model. AUROC and AUPRC was calculated excluding a feature at a time. Age had the greatest impact on model performance, followed by waist circumference, BMI, and total cholesterol.

**Figure 5.**
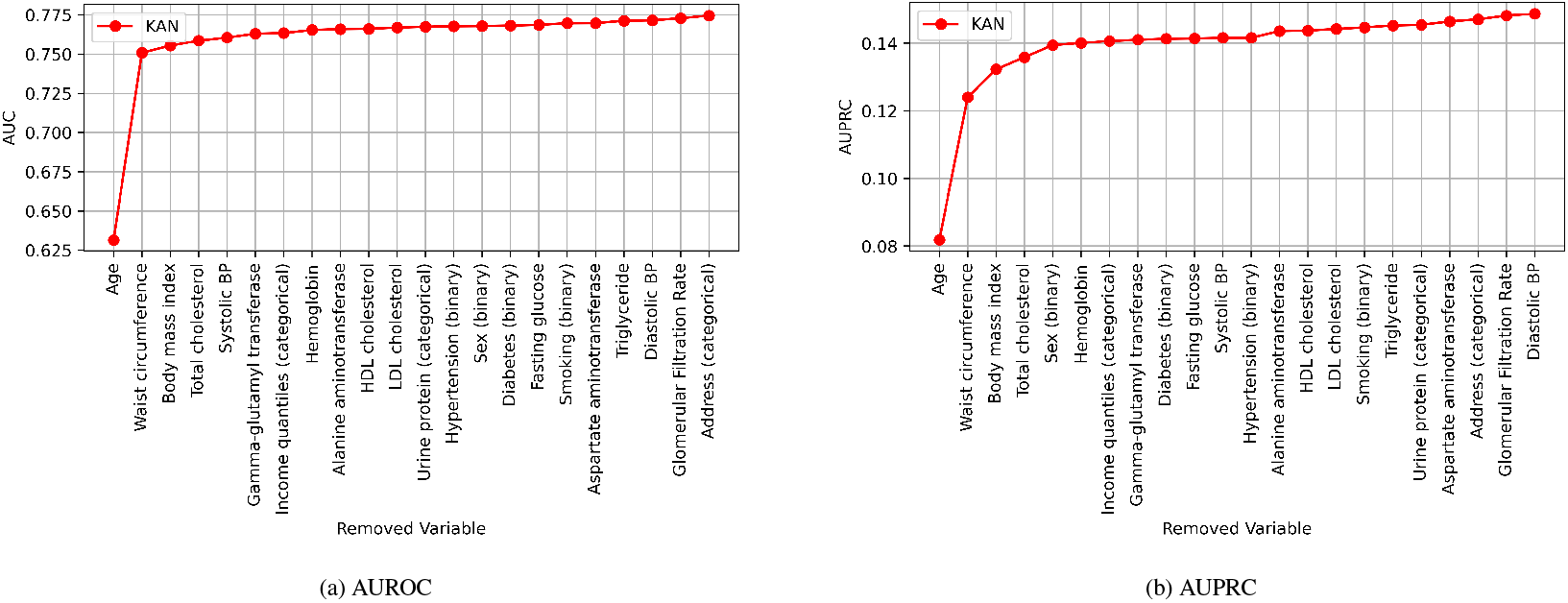
Performance on the test dataset after removing one specific variable at a time measured by (A) the area under the receiver operating characteristic (AUROC) and (B) the area under the precision recall curve (AUPRC)

## Discussion

In this study, we employed KAN, a novel deep learning architecture, to predict the 10-year ASCVD risk in a population-based cohort. Our KAN-based model showed the best performance among contemporary linear regression-based risk prediction models and other machine learning methods including MLP. Another advantage was interpretability. We were able to express the network and the activation functions on the edges as a formula and to visualize in a network graph. Additionally, we explored the influence of baseline features on ASCVD risk by analyzing SHAP values, performing ablation studies, and plotting risk prediction curves based on the model output.

Inspired by Kolmogorov-Arnold theorem, KANs utilize non-linear, learnable edges, which may enhance prediction accuracy. This study further explored whether KANs could provide additional improvements in accuracy. However, the observed performance gaps were modest: AUROCs for KAN, MLP, logistic regression, PREVENT, and KRPM all fell within 0.005. This likely reflects the inherently linear nature of the ASCVD risk prediction task. Despite their ability to model non-linear relationships, the advantages of KANs and MLPs appear limited in this context, evidenced by generally comparable performance of logistic regression across studies including ours.^33,35^ This suggests that the linear nature of the ASCVD risk prediction problem may diminish the benefits typically gained from nonlinear modeling approaches. Additionally, a random forest model—a popular machine learning technique in the medical field—performed significantly worse than the other methods.

Some claim that KAN is simply a specialized form of MLP. Yu et al.^61^ mathematically compared the forward process of KAN and MLP and analyzed whether the functional difference between the two architectures is substantial. Their findings show that KAN outperforms MLP only in symbolic formula representation, while MLP performs better in other tasks. Similary, Shukla et al.^62^ also conducted a comparative study to evaluate the potential and effectiveness of KAN-based representations to show that MLP outperforms KAN in all tasks except symbolic modeling. Further research is needed to determine which architecture provides greater accuracy across diverse domains. Nonetheless, KAN’s interpretability offers a significant advantage in the medical field by addressing the “black box” issue commonly raised with AI models. Unlike other machine learning models, KANs facilitate a more intuitive exploration of the effects of risk factors, as they can be expressed in human-readable formulas.

Our study has several limitations stemming from the characteristics of the data. The study cohort consisted of individuals who voluntarily participated in the national health screening program, which may lead to selection bias. Misclassification bias may also exist due to operation definition for key covariates and outcomes relying on claims information.^63^ Generalizability of this study may be limited, since this study was conducted exclusively in Korean population. However, to our knowledge, this is the first study to apply KAN to the 10-year ASCVD prediction task. Thus, rather than being a simple population-specific analysis, our work serves as a reference that such modeling can be extended to other populations.

Our KAN-based prediction model demonstrated superior performance compared to other machine learning models and conventional equation-based models. The model showed advantages in model interpretability. The network was able to be intuitively visualized and expressed in interpretable symbolic formulas. Analyses to understand the contribution of individual risk factors to the model output were also presented. In conclusion, KAN may be considered a machine learning model that can replace MLP in the medical domain by providing improved accuracy and interpretability.

## Data Availability

The data reported in this study are available to qualified researchers upon application to the National Health Insurance Sharing Service (https://nhiss.nhis.or.kr/) for the purposes of reproducing the results or replicating the procedures described in this study.

## Contributors

SK.K., SH.Kim and SH.Kang conceived of the presented idea. SH.Kim performed statistical analysis. SK.K. and W.K. contributed to development of the models. SH.Kim verified the analytical methods. CH.Y., TJ.Y. and IH.C. criticially reviewed the manuscript. All authors discussed the results and contributed to the final manuscript.

## Ethics statement

The Seoul National University Bundang Hospital’s institutional review board determined that our study was exempt from review (study number: X-2312-871-901).

## Acknowledgments

This research was supported by the Bio&Medical Technology Development Program of the National Research Foundation (NRF) funded by the Korean government (MSIT) (NO. RS-2023-00222910 and RS-2025-00517929).

## Human and Animal Rights and Informed Consent

The present study was performed in accordance with the Declaration of Helsinki and the need for informed consent was waived.

## Appendix A Machine learning models: MLP and KAN

### Appendix A.1 Multi-layer perceptron

The multi-layer perceptron, *f* ^MLP^, consists of *L* fully connected layers defined as ℱ_ℓ_ = *σ*(ℱ _ℓ−1_**w**_ℓ_ + *b*_ℓ_), ℓ = 1, …, *L*, where ℱ_0_ = **x**_*i*_ *∈* ℝ^*D*^, *i* = 1, …, *N* and **w**_1_ *∈* ℝ^*D*^, **w**_ℓ_ *∈* ℝ^*h*^, *b* ∈ ℝ are the learnable parameters (with *h* being the number of nodes in each hidden layer). The activation function *σ*(*·*) is a fixed, differentiable non-linear function applied element-wise. The final output of the MLP is given by

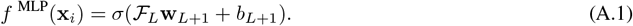

We selected the ReLU function as the nonlinear activation function, and *b* is defined as a scalar value representing the network bias.

### Appendix A.2 Kolmogorov-arnold network

We consider the problem of supervised learning on 𝒟, where the goal is to find a function *f* ^KAN^ such that *y ≈ f* ^KAN^(**x**) for inputs **x** *∈* ℝ^*D*^. Assuming that KAN is composed of *L* layers, its architecture can be represented as [*n*_0_, *n*_1_, …, *n*_*L*_], where *n*_ℓ_ *∈* ℤ+ denotes the number of nodes in layer for ℓ = 1, 2, …, *L*. For example, if the architecture is specified as [2, 5, 2], the input dimension is 2, the network contains a hidden layer with 5 nodes, and the output dimension is 2. To generalize, let *n*_ℓ_ denote the number of nodes in the ℓ-th layer. The *i*-th node in layer ℓ is indexed as (ℓ, *i*), and its activation function value is denoted by *x*_ℓ,*i*_. The number of activation functions between the ℓ-th and (ℓ + 1)-th layers is defined as *n*_ℓ_*n*_ℓ+1_. Each activation function connecting these two layers can be defined as follows:

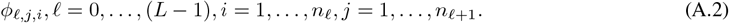

In this setting, *ϕ* is a learnable activation function. For a more detailed description, refer to the main body of the KAN formulation.^19^ Also *x*_ℓ+1,*j*_ can be defined as follows:

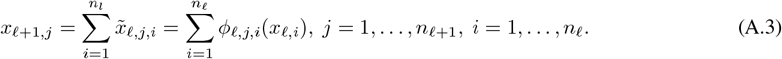

In accordance with the previously defined prediction model *f* : *→* [0, 1]^2^, we define a KAN with output dimension 2 and architecture [2,5,2] as follows:

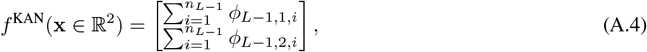

where *x*_ℓ,*i*_ serves as the input to *ϕ*_ℓ,*j,i*_. The detailed mathematical formulation of the KAN architecture can be found in Chapter 2 of origin paper.^19^

## Appendix B Scaling method

We applied the `StandardScaler` from scikit-learn,^51^ fitted on the training data, to transform all test inputs. The scaled values, denoted as **z**, are computed from the random variable vector **x** as follows:

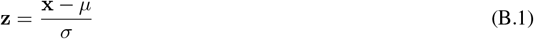

Where *µ* is the mean of each risk factor *x* computed from the training dataset, and *σ* is the corresponding standard deviation. The complete list of mean and standard deviation values is provided in Table B.1.

**Table B11:**
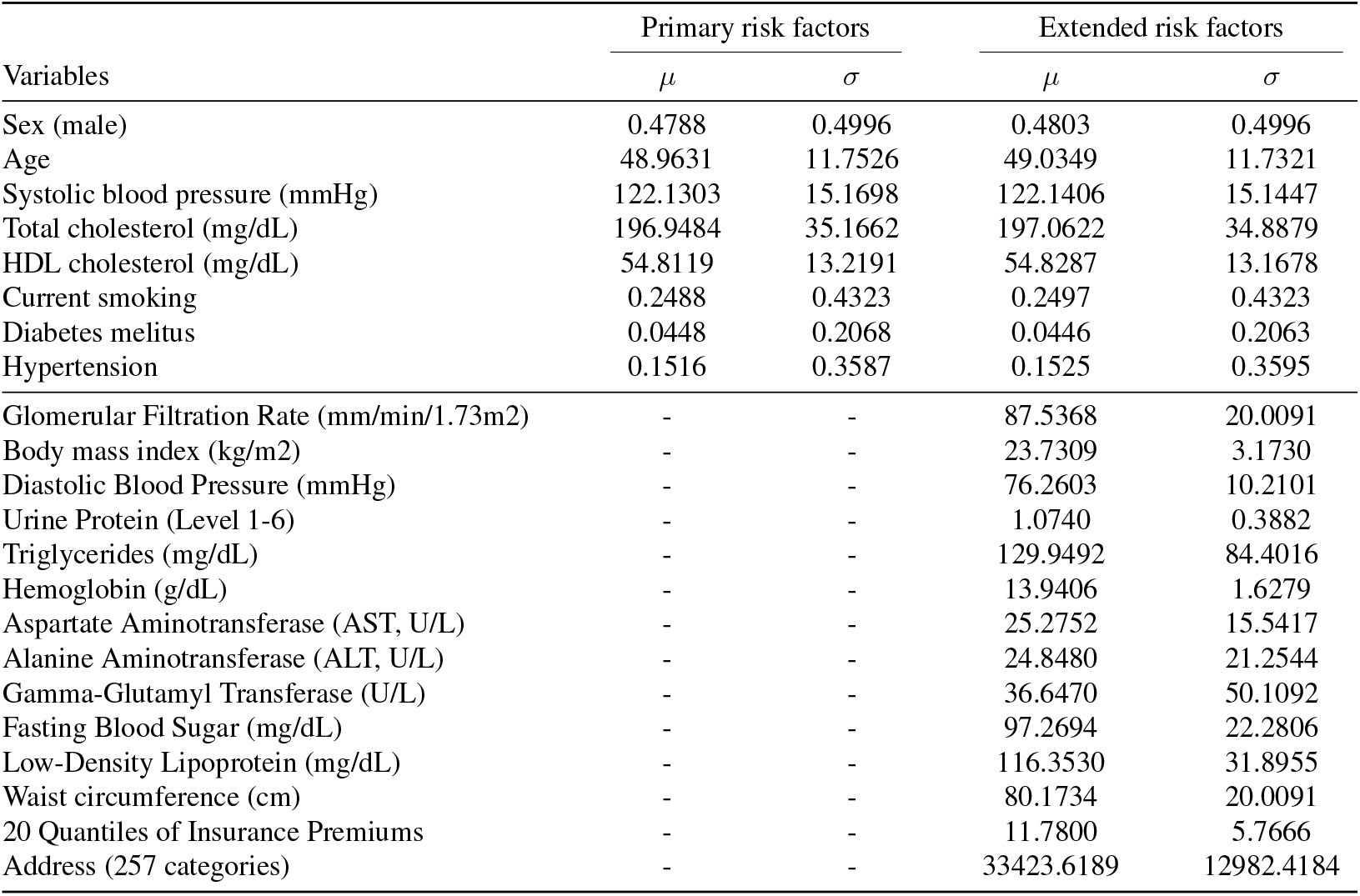
Mean and standard deviation for each variable set. Minor variations in overlapping variables, from sex to hypertension, arise due to differences in sample size after outlier filtering.

## Appendix C Primary KAN model

Unlike the extended model, the Primary KAN computes ASCVD risk using only eight input risk factors. The most compact form of the proposed Primary KAN consists of a single weighted summation term and an additional term involving a sine function, and is defined as follows.

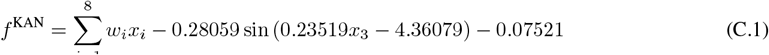

In the first term of Equation (C.1), the weights *w*_*d*_ and their corresponding input values *x*_*d*_, for *d* = 1, …, 8, are computed as follows: −0.0376*x*_1_(sex), 0.19224*x*_2_(age), −0.00877*x*_3_(systolic BP), 0.05861*x*_4_(current smoking), 0.01255*x*_5_(total cholesterol), −0.03263*x*_6_(hdl cholesterol), 0.03238*x*_7_(hypertension) and 0.04173*x*_8_(diabetes).

## Appendix D Figures

**Figure D.1:**
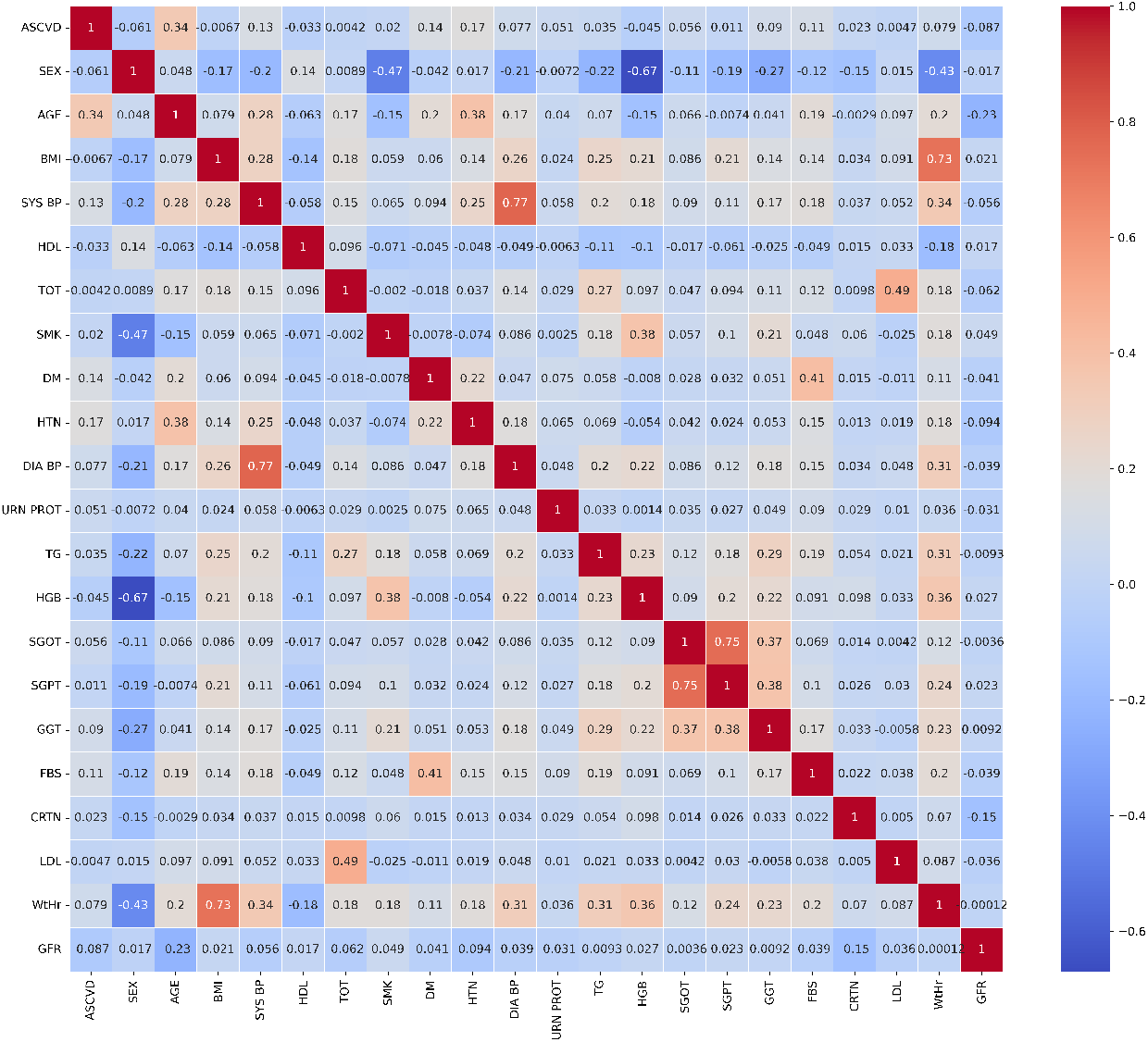
Correlation heatmap

**Figure D.2:**
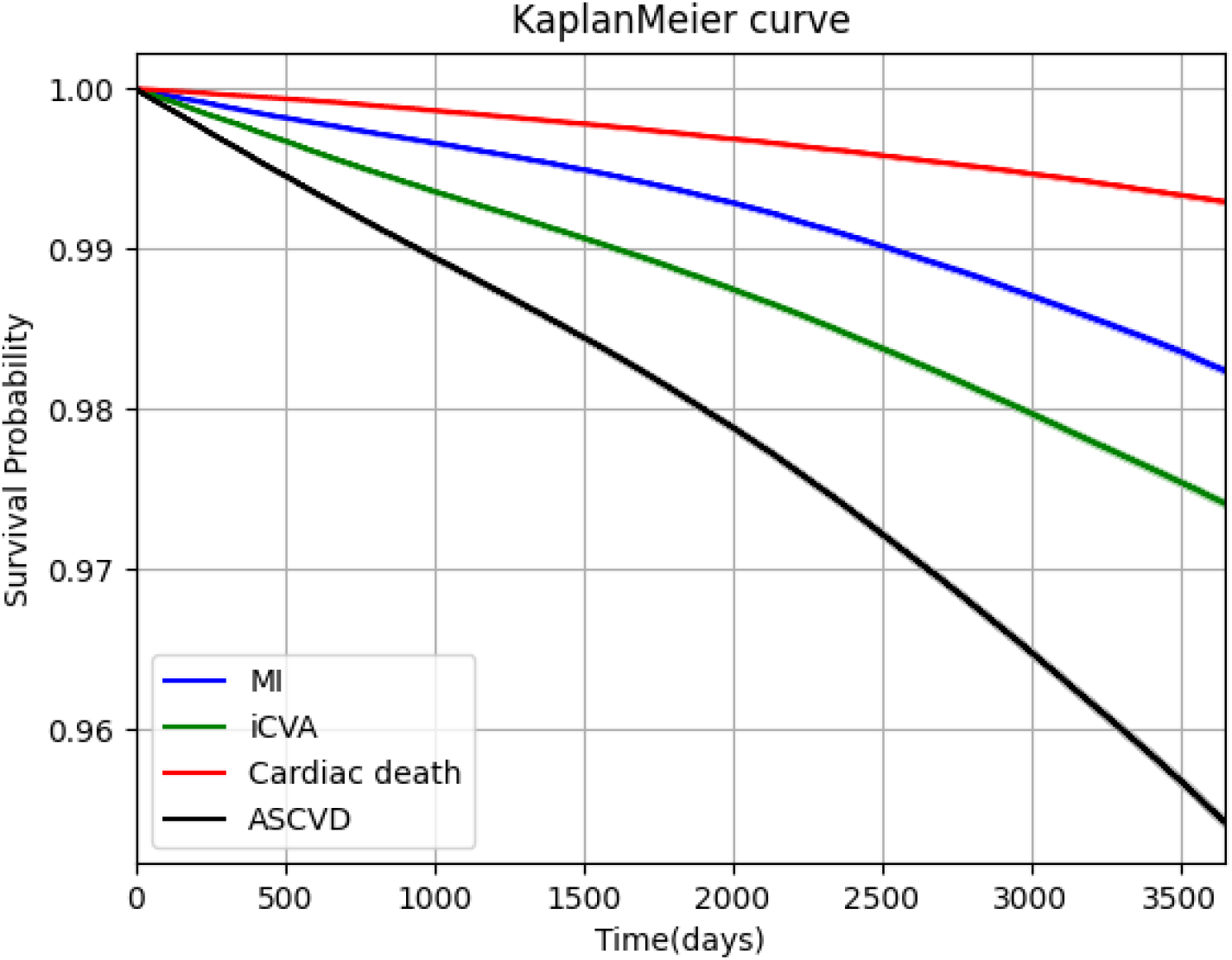
Kaplan meier curves

**Figure D.3:**
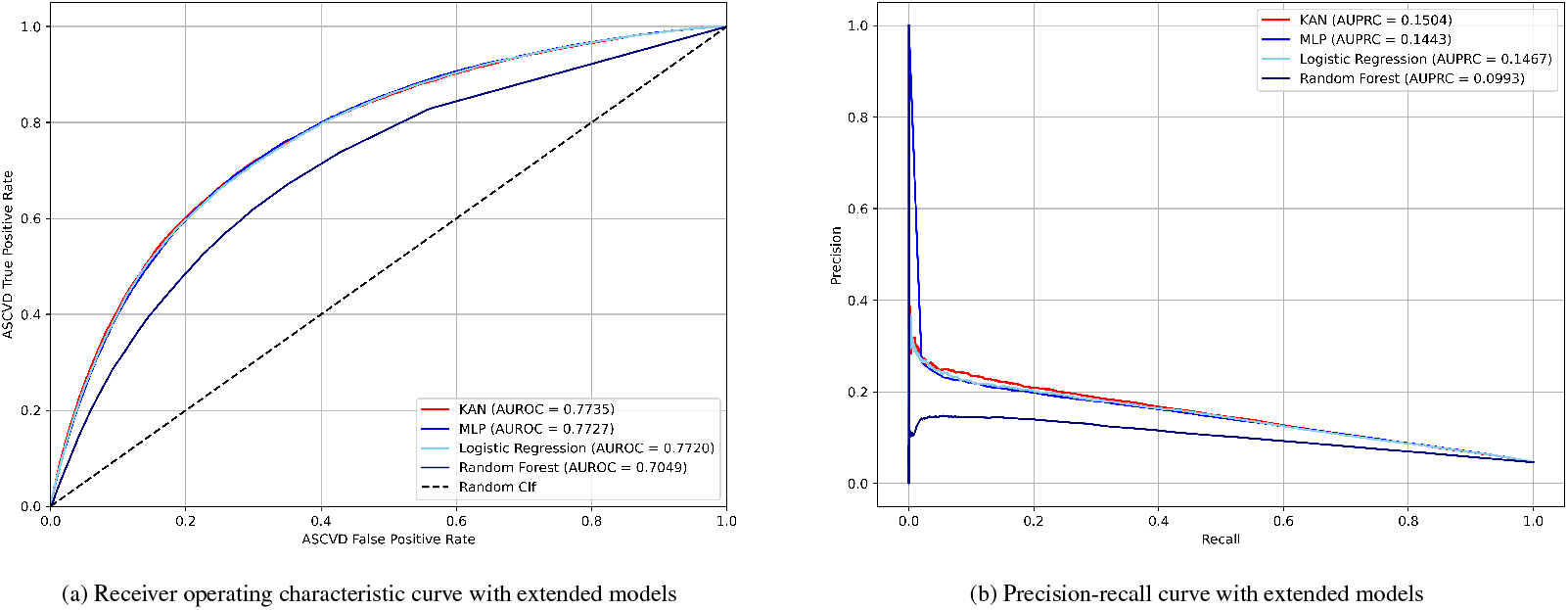
ROC curves and Precision-Recall curves for the primary models. Each curve illustrates the classification performance of a model across different thresholds. AUROC and AUPRC values are indicated in the legends for comparison.

## Appendix E Tables

**Table E.1:** Minimum and maximum thresholds used for outlier filtering. The thresholds were intentionally set leniently to exclude only a small number of data points.

|  | Minimum values | Maximum values |
| --- | --- | --- |
| **Variables in the primary model** |  |  |
| Sex (male) | 0 | 1 |
| Age | 30 | 79 |
| Systolic blood pressure (mmHg) | 70 | 250 |
| Total cholesterol (mg/dL) | 100 | 400 |
| HDL cholesterol (mg/dL) | 20 | 100 |
| Current smoking | 0 | 1 |
| Diabetes mellitus | 0 | 1 |
| Hypertension | 0 | 1 |
| **Variables in the extended model** |  |  |
| Glomerular Filtration Rate (mm/min/1.73m<sup>2</sup>) | 0 | 250 |
| Body mass index (kg/m<sup>2</sup>) | 5 | 70 |
| Diastolic Blood Pressure (mmHg) | 40 | 150 |
| Urine Protein (Level 1-6) | 0 | 6 |
| Triglycerides (mg/dL) | 30 | 1000 |
| Hemoglobin (g/dL) | 6 | 20 |
| Aspartate Aminotransferase (AST, U/L) | 0 | 1000 |
| Alanine Aminotransferase (ALT, U/L) | 0 | 1000 |
| Gamma-Glutamyl Transferase (U/L) | 0 | 1000 |
| Fasting Blood Sugar (mg/dL) | 30 | 500 |
| Low-Density Lipoprotein (mg/dL) | 30 | 300 |
| Waist circumference (cm) | 30 | 250 |
| 20 Quantiles of Insurance Premiums | 0 | 20 |
| Address (257 categories) | 11110 | 50130 |

**Table E.2:** Variables urine protein level and quantiles of insurance premiums extracted from the Korean National Health Insurance Service Cohort dataset.

| Categorical risk factors | Number (Proportion) |
| --- | --- |
| **Urine Protein Level** |  |
| Level 1 | 2,015,323 / 2,116,621 (95.21%) |
| Level 2 | 46,875 / 2,116,621 (2.21%) |
| Level 3 | 31,280 / 2,116,621 (1.48%) |
| Level 4 | 11,050 / 2,116,621 (0.52%) |
| Level 5 | 2,889 / 2,116,621 (0.14%) |
| Level 6 | 542 / 2,116,621 (0.03%) |
| **Quantiles of Insurance Premiums** |  |
| 1 | 14,810 / 2,116,621 (0.7%) |
| 2 | 74,650 / 2,116,621 (3.53%) |
| 3 | 75,622 / 2,116,621 (3.57%) |
| 4 | 78,395 / 2,116,621 (3.7%) |
| 5 | 69,749 / 2,116,621 (3.3%) |
| 6 | 77,444 / 2,116,621 (3.66%) |
| 7 | 81,617 / 2,116,621 (3.86%) |
| 8 | 73,031 / 2,116,621 (3.45%) |
| 9 | 84,457 / 2,116,621 (3.99%) |
| 10 | 87,537 / 2,116,621 (4.14%) |
| 11 | 95,480 / 2,116,621 (4.51%) |
| 12 | 98,276 / 2,116,621 (4.64%) |
| 13 | 106,435 / 2,116,621 (5.03%) |
| 14 | 109,355 / 2,116,621 (5.17%) |
| 15 | 115,915 / 2,116,621 (5.48%) |
| 16 | 124,841 / 2,116,621 (5.9%) |
| 17 | 131,833 / 2,116,621 (6.23%) |
| 18 | 137,594 / 2,116,621 (6.5%) |
| 19 | 142,561 / 2,116,621 (6.74%) |
| 20 | 144,253 / 2,116,621 (6.82%) |

**Table E.3:**
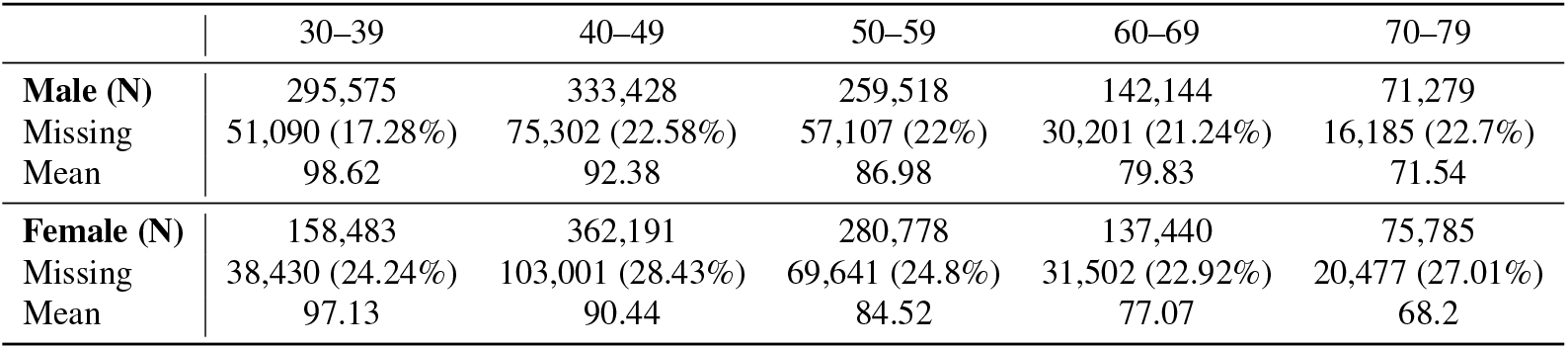
eGFR imputation information.

|  | 30–39 | 40–49 | 50–59 | 60–69 | 70–79 |
| --- | --- | --- | --- | --- | --- |
| **Male (N)** | 295,575 | 333,428 | 259,518 | 142,144 | 71,279 |
| Missing | 51,090 (17.28%) | 75,302 (22.58%) | 57,107 (22%) | 30,201 (21.24%) | 16,185 (22.7%) |
| Mean | 98.62 | 92.38 | 86.98 | 79.83 | 71.54 |
| **Female (N)** | 158,483 | 362,191 | 280,778 | 137,440 | 75,785 |
| Missing | 38,430 (24.24%) | 103,001 (28.43%) | 69,641 (24.8%) | 31,502 (22.92%) | 20,477 (27.01%) |
| Mean | 97.13 | 90.44 | 84.52 | 77.07 | 68.2 |

## Footnotes

1 Unlike the definition in Equation (1), logistic regression was implemented to produce a single output following the conventional approach.

2 Confidence interval

3 A simpler version, the primary KAN model is provided in the Appendix C.

